# Validation of the Enhanced Recovery After Surgery (ERAS) database in Alberta, Canada

**DOI:** 10.1101/2025.03.01.25322874

**Authors:** K Sauro, A Thomas, L Bakunda, C Smith, S Ibadin, T Kuzma, G Nelson

## Abstract

**Background:** The Enhanced Recovery After Surgery (ERAS) Interactive Audit System (EIAS) is a retrospectively collected database containing information about the preoperative, intraoperative, and postoperative components of surgical patient care. EIAS was created to allow centers that have adopted ERAS protocols to assess their performance. To have confidence in the data collected by EIAS, its completeness, accuracy and validity must be assessed. This study aims to assess the validity of the Alberta EIAS when compared to the gold standard measurement for patient data, the patient electronic medical record (EMR).

**Methods:** Four sites that implemented ERAS across Alberta were included, with 20 to 60 patient EMRs pulled from each site. Data on fourteen pre-specified variables was abstracted from patient EMRs and compared to the corresponding variables from EIAS. Validation criteria included (I) accuracy (agreement between EMR and EIAS) and (II) missingness (percent of data that was missing in patients EMR and EIAS). The estimates of accuracy were compared to estimates of accuracy from two other EIAS validation studies using meta-analysis.

**Results:** A total of 113 patient charts were reviewed across four sites. Agreement between chart review and EIAS was 73.6% (59.9% - 87.3%) with a mean sensitivity of 70.3 and mean specificity of 50.1. Agreement between chart review and EIAS was better among outcomes (agreement for re-operation was 93.7%) than it was for accuracy of documentation of the ERAS elements (mean agreement=73.6%). Agreement varied by site (68.5% to 94.4%) and reviewer (68.0% to 96.6%). Across all fifteen ERAS elements, a mean of 11.4% of data was missing, with re-operation having the greatest proportion of missing data (15.9%) and termination of drains and early mobilization with the lowest proportion of missing data (9.7%). Estimates of accuracy were not different between studies (I^2^=56.4%, p=0.101).

**Conclusions:** In Alberta, EIAS is an accurate and complete source of data suggesting that EIAS is a valid and reliable source of data to explore patient outcomes and adoption of ERAS guidelines. This study found that data abstractors that are medically trained, and trained in standardized data abstraction are important determinants of generating high quality data, highlighting the need for adequate resources for data collection.

## INTRODUCTION

Enhanced Recovery After Surgery (ERAS) guidelines are an evidence-based approach to pre-, intra- and post-operative care that have been found to reduce complications, hospital length of stay without increasing hospital readmission, and costs.(1) The improvement in patient and healthcare system outcomes after implementing ERAS guidelines has led to increased reach of the guidelines across surgical specialties and across the world.(2) Due to the widespread use of ERAS across the globe, comparing implementation of ERAS guidelines (including compliance) and patient outcomes is important to better understand the effectiveness of ERAS.

Among the key components of ERAS is auditing clinical practice to assess adoption of ERAS guidelines. To this end the ERAS Interactive Audit System (EIAS) has been developed to streamline and standardize data collection for studies of ERAS effectiveness and compliance. The web-based EIAS collects data on approximately 150 variables related to the patient and perioperative care and provides dashboards to visualize care processes.(3) While EIAS has the potential to be a powerful tool for evaluating compliance with ERAS guidelines and the effectiveness of ERAS guidelines, as with any database, the quality of the findings are dependent on the quality of the data. Two studies have explored the quality of EIAS data, in Sweden and Switzerland.(4, 5) These studies have found that the agreement between medical charts and the data captured in EIAS is good and coverage was high, which provides confidence in the findings of studies emerging from the EIAS data in these centres.(4, 5) However, it is unclear if the quality of data from EIAS is similar globally, or beyond Europe.

The objective of this study was to compare the quality of the EIAS data from an ERAS Centre of Excellence in Canada(6) to the data quality in these previous papers.(4, 5) The quality of the data from Alberta, Canada was evaluated and the estimates from all three studies were pooled to explore heterogeneity between EIAS data quality globally.

## METHODS

### Design & Setting

This retrospective cohort study was conducted in Alberta, Canada which is an ERAS Centre of Excellence.(6) Alberta is the fourth most populated province in Canada, with a population of 4.3 million. Canada is a universal, publicly funded healthcare system where health services are delivered by the provinces. In Alberta, there is a single health services provider, Alberta Health Services, which implemented ERAS across the province in 2013. Initially, ERAS was implemented for colorectal surgery in the main urban, tertiary care hospitals but has expanded to 9 hospitals across the province.(7) The implementation of ERAS in Alberta included the integration of EIAS in January 2013.

This study was approved by the University of Calgary Conjoint Health Research Ethics Board (REB21-1021).

### Participants

Adult patients with a record in EIAS from January 2013 to December 2022 were included. A random sample of 20 patients at four hospitals (n=113) were selected to evaluate the accuracy of the data in EIAS compared to the medical charts, except the largest hospital in the province (Foothills Medical Centre, Calgary, Alberta) was over sampled (n=60); a stratified random sample of 10 patients for each of the six types of ERAS-guided surgeries fully implemented were selected from this site.

### Outcomes

Two aspects of data quality: accuracy and missingness were evaluated.

*Accuracy* refers to how accurately the EIAS data entry was completed. To evaluate the accuracy of data for each of twelve common components of ERAS (preoperative anemia, preoperative nutrition, avoidance of oral bowel preparation, intraoperative antibiotics, intraoperative normothermia, postoperative nausea and vomiting prophylaxis, avoidance of nasogastric tube use, venous thromboembolism prophylaxis, termination of urinary drainage, postoperative euvolemia, early feeding, and early mobilization) and three patient outcomes (re-operations, readmissions, and complications), included in EIAS, a binary “present” or “not present” was abstracted from the chart; if the data element was discussed in the medical chart then it was deemed “present” but if was not noted in the chart then it was “not present.” The standardized data abstraction form is available in Supplemental File 1. Concordance between EIAS and electronic medical records on hospital readmission, complications using the Clavien-Dindo classification and reoperation were also evaluated. The percent agreement and kappa score between EIAS and medical record was calculated.

*Missingness* refers to the proportion of variables missing for the primary outcome variable, compliance and the outcome variables evaluated for accuracy. The same patient records used to evaluate accuracy were used to evaluate missingness. To determine if missing values were missing at random or if they were due to systematic error, a Fisher’s exact test was used to determine if missingness was related to six patient and hospital-level factors (age, sex, hospital size, hospital type, season of admission, surgery type).

### Data Collection

Data abstraction from the medical charts was done using a standardized data abstraction form. Five reviewers were trained to abstract the data from the medical charts; one reviewer was a fellow in a surgical training program, one was an internationally trained physician in public health, one was a PhD trained health services researcher, and the remaining two reviewers were research assistants trained to conduct the chart review.

To assess missingness, a population-based administrative dataset was used. The Discharge Abstract Database (DAD) is a federally mandated database that collects information on hospital stay using a standardized data abstraction process by trained coders.(8) The DAD data was provided by the single data custodian for Alberta, AHS and was deterministically linked to the EIAS using a personal health number assigned to each resident of Alberta at birth or upon immigration to the province, which follows them throughout their life.

### Analysis

Accuracy between medical charts and EIAS was assessed for three patient outcomes and twelve ERAS elements by calculating the percent agreement (true positives and true negatives divided by the total number of patients, using medical chart data as the gold standard). Agreement was stratified by reviewer and hospital site. Sensitivity (number of patients with data for the variable present in their medical chart and EIAS divided by the number of patients with data present in their medical chart) and specificity (number of patients without data present for the variable in their medical chart and EIAS divided by the number of patients without data for the variable present in their medical chart) were calculated using medical chart data as the gold standard.

Missingness was estimated by calculating the percentage of missing data for each variable for the entire cohort, as well as by hospital site and reviewer.

Random-effects meta-analysis was conducted to pool estimates of mean accuracy (using the metan command).

STATA version 18.5 was used for all analyses.

## RESULTS

### Participants

113 patients were included in the study. Most commonly patients had colorectal surgery (n=67, 59%) and were treated at Foothills Medical Centre (n=52, 46%). Patients were admitted to hospital between January 2017 and March 2022.

### Accuracy

Overall, agreement between medical charts and EIAS was 73.6% (59.9% - 87.3%) with a mean sensitivity of 70.3 and mean specificity of 50.1 (Table 1). Agreement varied by the outcome and ERAS element assessed, with the highest agreement for re-operation, preoperative anemia, and early mobilization and the worst agreement for preoperative nutrition, avoidance of oral bowel preparation, and avoidance of nasogastric tube use (Table 1). Sensitivity and specificity varied by outcome and ERAS element, with termination of urinary drainage, early mobilization, and postoperative nausea and vomiting prophylaxis having the highest sensitivity, and preoperative anemia, intraoperative normothermia, and early feeding having the highest specificity (Table 1). Agreement also varied by site and reviewer. The highest mean agreement was among patients admitted to the Chinook Regional Hospital (94.4%) while the Red Deer Regional Hospital had the lowest mean agreement at 68.5% (Table 2). Agreement also varied by reviewer, with mean agreement ranging from 68.0% to 96.6% across all five reviewers (Table 2).

**Table 1:** Accuracy of EIAS data compared to chart review across all sites and reviewers.

|  | TP | FN | FP | TN | Sensitivity | Specificity | Agreement | Kappa |
| --- | --- | --- | --- | --- | --- | --- | --- | --- |
| **Patient Outcomes** |  |  |  |  |  |  |  |  |
| Re-operations | 1 | 4 | 3 | 103 | 20.00 | 97.17 | 93.69 | 0.1898 |
| Re-admissions | 4 | 3 | 13 | 91 | 57.15 | 87.50 | 85.59 | 0.2679 |
| Complications | 16 | 3 | 24 | 32 | 84.20 | 57.1 | 64.00 | 0.3029 |
| **ERAS Elements** |  |  |  |  |  |  |  |  |
| Preoperative Anemia | 0 | 7 | 0 | 104 | 0.00 | 100.00 | 93.69 | 0.0000 |
| Preoperative Nutrition | 42 | 2 | 61 | 6 | 95.45 | 8.96 | 43.24 | 0.0358 |
| Oral Bowel Preparation | 37 | 1 | 49 | 24 | 97.37 | 32.88 | 54.95 | 0.2322 |
| Intraoperative Antibiotics | 75 | 2 | 30 | 4 | 97.40 | 11.77 | 71.17 | 0.1190 |
| Intraoperative Normothermia | 19 | 19 | 0 | 73 | 50.00 | 100.00 | 82.88 | 0.0856 |
| PONV Prophylaxis | 66 | 1 | 38 | 6 | 98.51 | 13.64 | 64.86 | 0.1419 |
| Nasogastric Tube | 51 | 9 | 34 | 17 | 85.00 | 33.34 | 61.26 | 0.1903 |
| VTE Prophylaxis | 77 | 13 | 19 | 2 | 85.56 | 9.53 | 71.17 | 0.0000 |
| Termination of Urinary Drainage | 85 | 0 | 26 | 0 | 100.00 | 0.00 | 76.58 | 0.0000 |
| Postoperative Euvolemia | 19 | 32 | 0 | 60 | 37.25 | 100.00 | 71.17 | 0.3909 |
| Early Mobilization | 99 | 0 | 12 | 0 | 100.00 | 0.00 | 89.19 | 0.0000 |
| Early Feeding | 19 | 22 | 0 | 70 | 46.43 | 100.00 | 80.18 | 0.5214 |
| **Overall (Mean)** |  |  |  |  | 70.29 | 50.13 | 73.6 |  |

*Abbreviations:* TP = true positive; FN = false negative; FP = false positive; TN = true negative; PONV = postoperative nausea and vomiting; VTE = venous thromboembolism.

**Table 2:**
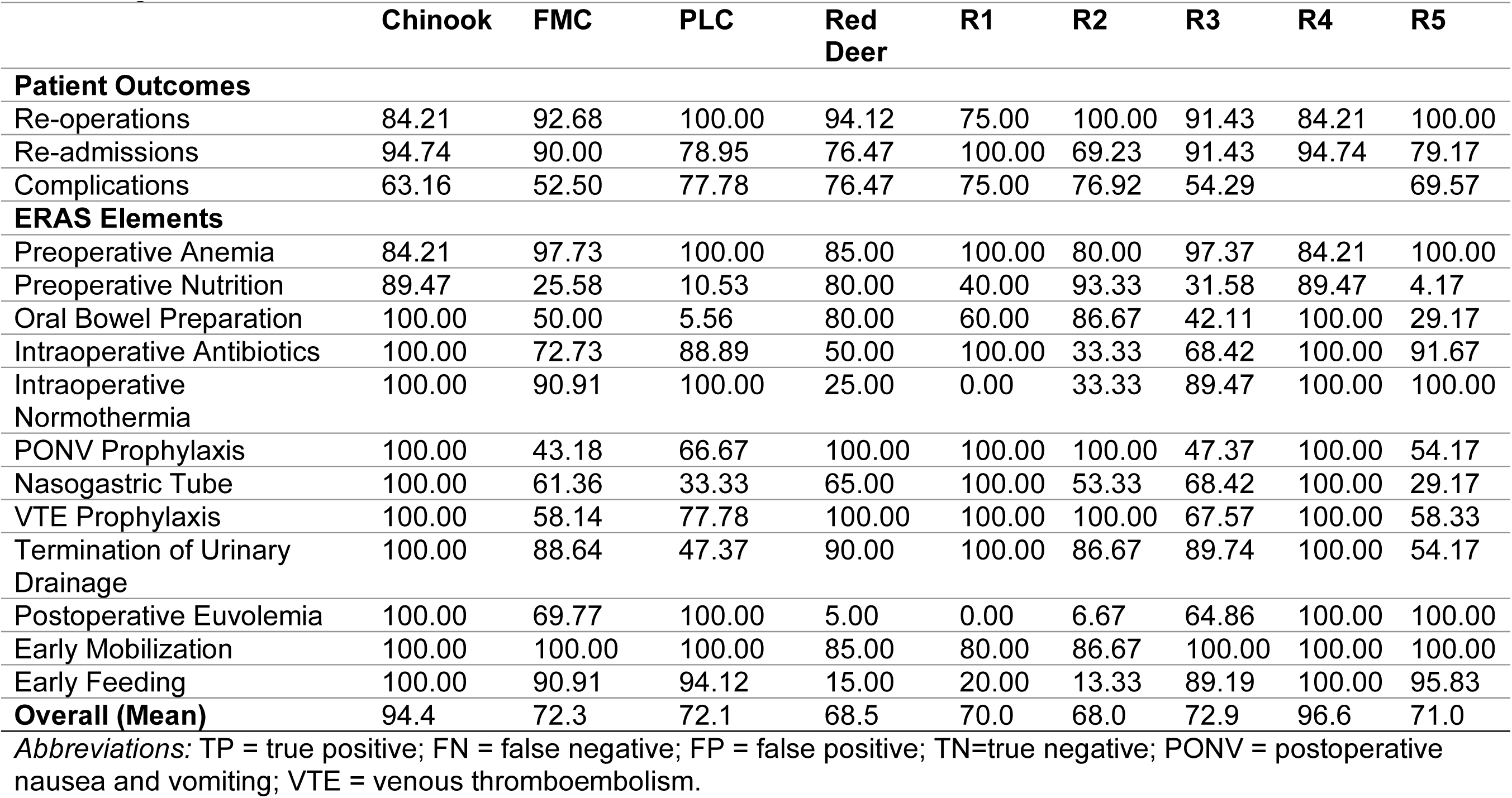
Agreement across sites and reviewers.

|  | **Chinook** | **FMC** | **PLC** | **Red Deer** | **R1** | **R2** | **R3** | **R4** | **R5** |
| --- | --- | --- | --- | --- | --- | --- | --- | --- | --- |
| **Patient Outcomes** |  |  |  |  |  |  |  |  |  |
| Re-operations | 84.21 | 92.68 | 100.00 | 94.12 | 75.00 | 100.00 | 91.43 | 84.21 | 100.00 |
| Re-admissions | 94.74 | 90.00 | 78.95 | 76.47 | 100.00 | 69.23 | 91.43 | 94.74 | 79.17 |
| Complications | 63.16 | 52.50 | 77.78 | 76.47 | 75.00 | 76.92 | 54.29 |  | 69.57 |
| **ERAS Elements** |  |  |  |  |  |  |  |  |  |
| Preoperative Anemia | 84.21 | 97.73 | 100.00 | 85.00 | 100.00 | 80.00 | 97.37 | 84.21 | 100.00 |
| Preoperative Nutrition | 89.47 | 25.58 | 10.53 | 80.00 | 40.00 | 93.33 | 31.58 | 89.47 | 4.17 |
| Oral Bowel Preparation | 100.00 | 50.00 | 5.56 | 80.00 | 60.00 | 86.67 | 42.11 | 100.00 | 29.17 |
| Intraoperative Antibiotics | 100.00 | 72.73 | 88.89 | 50.00 | 100.00 | 33.33 | 68.42 | 100.00 | 91.67 |
| Intraoperative Normothermia | 100.00 | 90.91 | 100.00 | 25.00 | 0.00 | 33.33 | 89.47 | 100.00 | 100.00 |
| PONV Prophylaxis | 100.00 | 43.18 | 66.67 | 100.00 | 100.00 | 100.00 | 47.37 | 100.00 | 54.17 |
| Nasogastric Tube | 100.00 | 61.36 | 33.33 | 65.00 | 100.00 | 53.33 | 68.42 | 100.00 | 29.17 |
| VTE Prophylaxis | 100.00 | 58.14 | 77.78 | 100.00 | 100.00 | 100.00 | 67.57 | 100.00 | 58.33 |
| Termination of Urinary Drainage | 100.00 | 88.64 | 47.37 | 90.00 | 100.00 | 86.67 | 89.74 | 100.00 | 54.17 |
| Postoperative Euvolemia | 100.00 | 69.77 | 100.00 | 5.00 | 0.00 | 6.67 | 64.86 | 100.00 | 100.00 |
| Early Mobilization | 100.00 | 100.00 | 100.00 | 85.00 | 80.00 | 86.67 | 100.00 | 100.00 | 100.00 |
| Early Feeding | 100.00 | 90.91 | 94.12 | 15.00 | 20.00 | 13.33 | 89.19 | 100.00 | 95.83 |
| **Overall (Mean)** | 94.4 | 72.3 | 72.1 | 68.5 | 70.0 | 68.0 | 72.9 | 96.6 | 71.0 |

*Abbreviations:* TP = true positive; FN = false negative; FP = false positive; TN=true negative; PONV = postoperative nausea and vomiting; VTE = venous thromboembolism.

Agreement varied by surgery, with the highest mean agreement among patients who received colorectal surgery (73.19%) while patients who underwent urologic surgeries had the lowest mean agreement at 69.13% (Supplemental File 2).

### Missingness

Across all 12 ERAS elements, a mean of 11.4% of data was missing. Re-operation and readmissions had the greatest proportion of missing data (15.9%), while termination of urinary drainage and early mobilization had the lowest proportion of missing data (9.7%) (Table 3). Across hospital sites, the mean percent of missing data was lowest at Chinook Regional Hospital (0.0%), and highest at Foothills Medical Centre (16.7%). Missing data across reviewers ranged from 0% to 8.3% (Table 3).

**Table 3:** Percentage of Missing Data for each Patient Outcome and ERAS Element, Stratified by Hospital Site and Reviewer.

|  | Overall | FMC | PLC | RDRH | CRH | R1 | R2 | R3 | R4 | R5 |
| --- | --- | --- | --- | --- | --- | --- | --- | --- | --- | --- |
| **Patient Outcomes** |  |  |  |  |  |  |  |  |  |  |
| Re-operations | 15.9 | 21.2 | 18.2 | 15.0 | 0 | 20.0 | 13.3 | 14.6 | 0 | 7.7 |
| Re-admissions | 15.9 | 23.0 | 13.7 | 15.0 | 0 | 20.0 | 13.3 | 14.6 | 0 | 7.7 |
| Complications | 10.6 | 15.4 | 4.5 | 15.0 | 0 | 20.0 | 13.3 | 4.9 | 0 | 0 |
| **ERAS Elements** |  |  |  |  |  |  |  |  |  |  |
| Preoperative Anemia | 10.6 | 15.4 | 18.2 | 0 | 0 | 0 | 0 | 7.3 | 0 | 7.7 |
| Preoperative Nutrition | 10.6 | 17.3 | 13.7 | 0 | 0 | 0 | 0 | 7.3 | 0 | 7.7 |
| Oral Bowel Preparation | 10.6 | 15.4 | 18.2 | 0 | 0 | 0 | 0 | 7.3 | 0 | 7.7 |
| Intraoperative Antibiotics | 10.6 | 15.4 | 18.2 | 0 | 0 | 0 | 0 | 7.3 | 0 | 7.7 |
| Intraoperative Normothermia | 10.6 | 15.4 | 18.2 | 0 | 0 | 0 | 0 | 7.3 | 0 | 7.7 |
| PONV Prophylaxis | 10.6 | 15.4 | 18.2 | 0 | 0 | 0 | 0 | 7.3 | 0 | 7.7 |
| Nasogastric Tube | 10.6 | 15.4 | 18.2 | 0 | 0 | 0 | 0 | 7.3 | 0 | 7.7 |
| VTE Prophylaxis | 12.4 | 17.3 | 18.2 | 5.0 | 0 | 0 | 6.7 | 9.8 | 0 | 7.7 |
| Termination of Urinary Drainage | 9.7 | 15.4 | 13.7 | 0 | 0 | 0 | 0 | 4.9 | 0 | 7.7 |
| Postoperative Euvolemia | 11.5 | 17.3 | 18.2 | 0 | 0 | 0 | 0 | 9.8 | 0 | 7.7 |
| Early Mobilization | 9.7 | 15.4 | 13.7 | 0 | 0 | 0 | 0 | 4.9 | 0 | 7.7 |
| Early Feeding | 11.5 | 15.4 | 22.7 | 0 | 0 | 0 | 0 | 9.8 | 0 | 7.7 |
| **Overall** | 11.4 | 16.7 | 16.4 | 3.3 | 0 | 4.0 | 3.1 | 8.3 | 0 | 7.2 |

### Pooled estimates of accuracy

The estimates from two studies; both described the accuracy of the EIAS database in patients undergoing colorectal surgery with one cohort representing surgical centres in Switzerland between January 2017 and December 2017,(4) and the other in Swedish centres.(5) When comparing the accuracy reported in the present study to other studies exploring accuracy, the accuracy reported in the present study was lower than that of other centres but was not significantly different (Figure 1). Similar trends in sensitivity and specificity were observed across centres (Figure 2).

**Figure 1:**
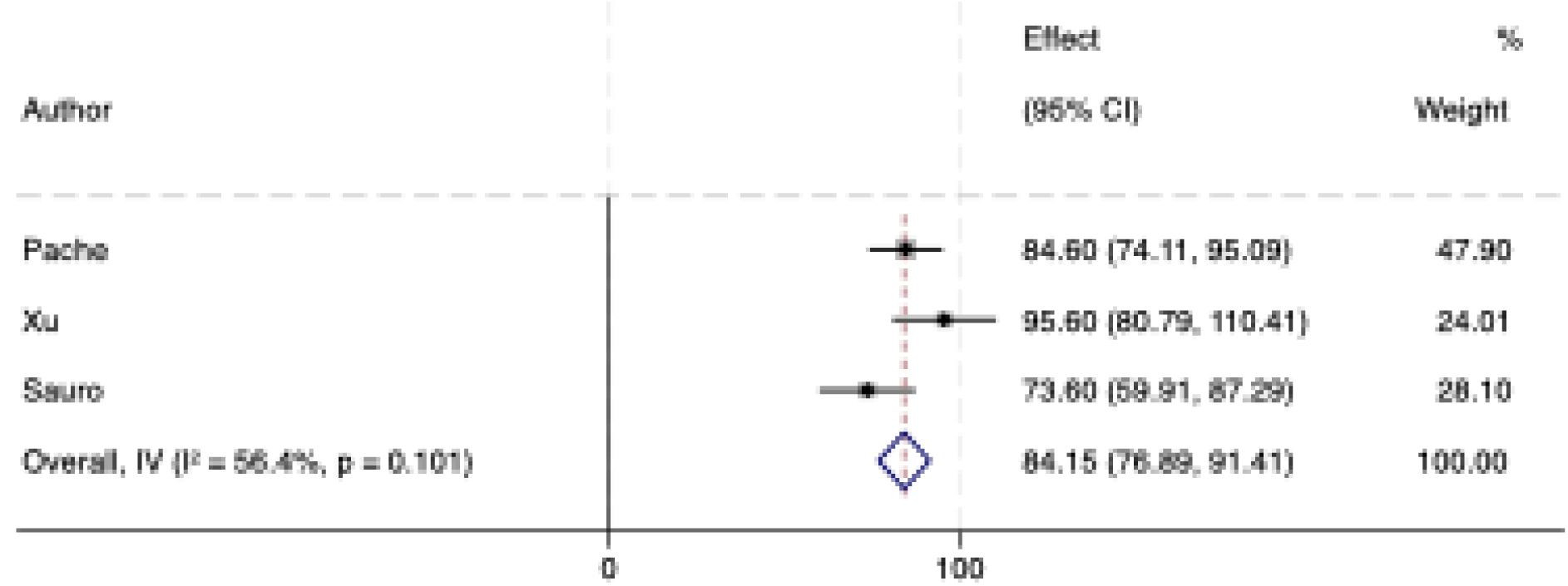
Forest plot of meta-analysis of accuracy. Note: Metan command used as mean accuracy across all variables, and standard error were used to for meta-analysis. Standard error was calculated if it was not reported.

**Figure 2:**
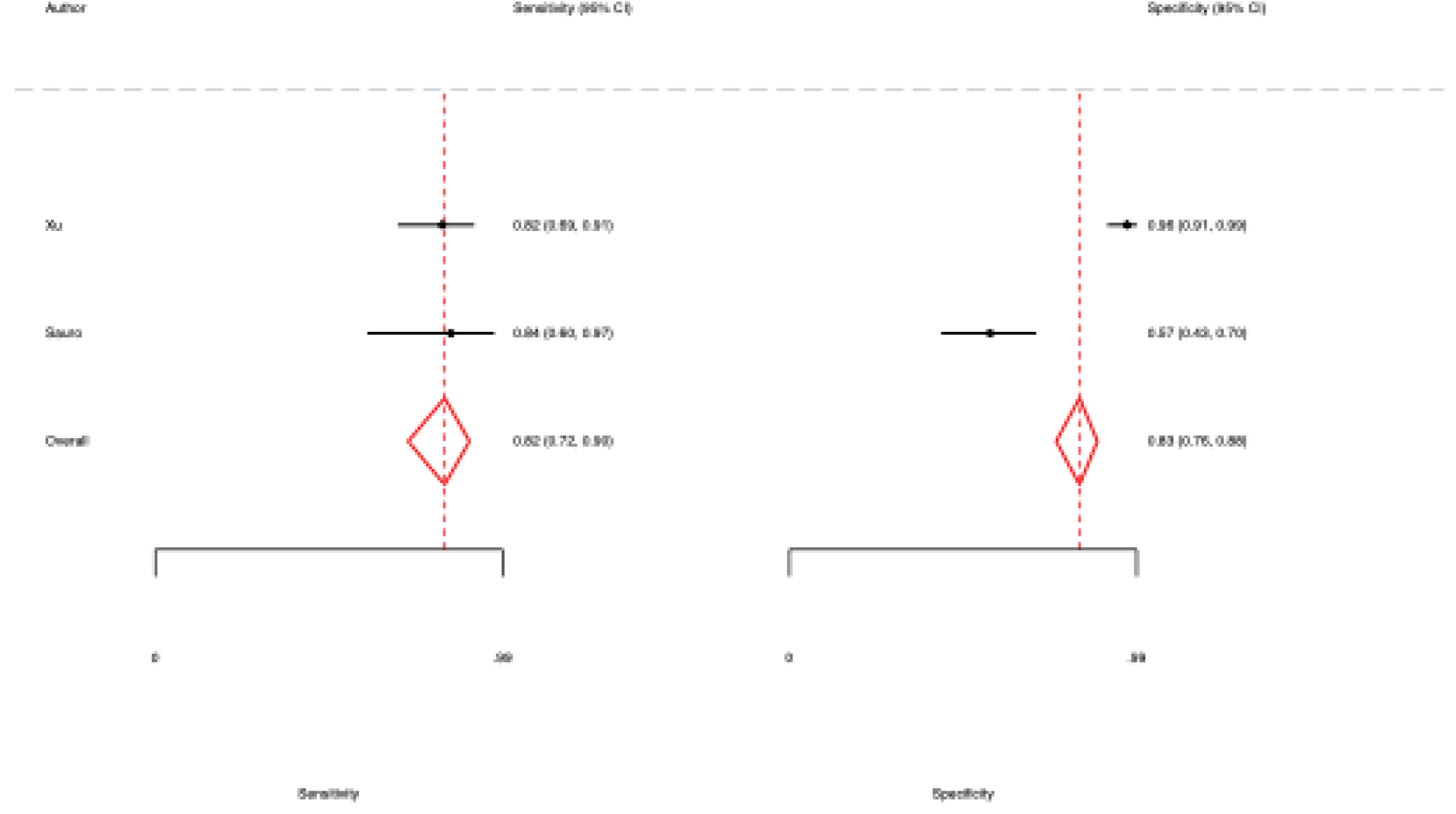
Forest plot of meta-analysis of measures of diagnostic accuracy for complications. *Notes:* Only Xu and Sauro are included in meta-analysis as Pache did not provide enough data in their manuscript and supplemental files to reconstruct the necessary two-by-two tables. The metadta command in Stata was used rather than metandi as metandi requires at least four studies. Fixed-effects model used as n < 3.

**Figure 3:**
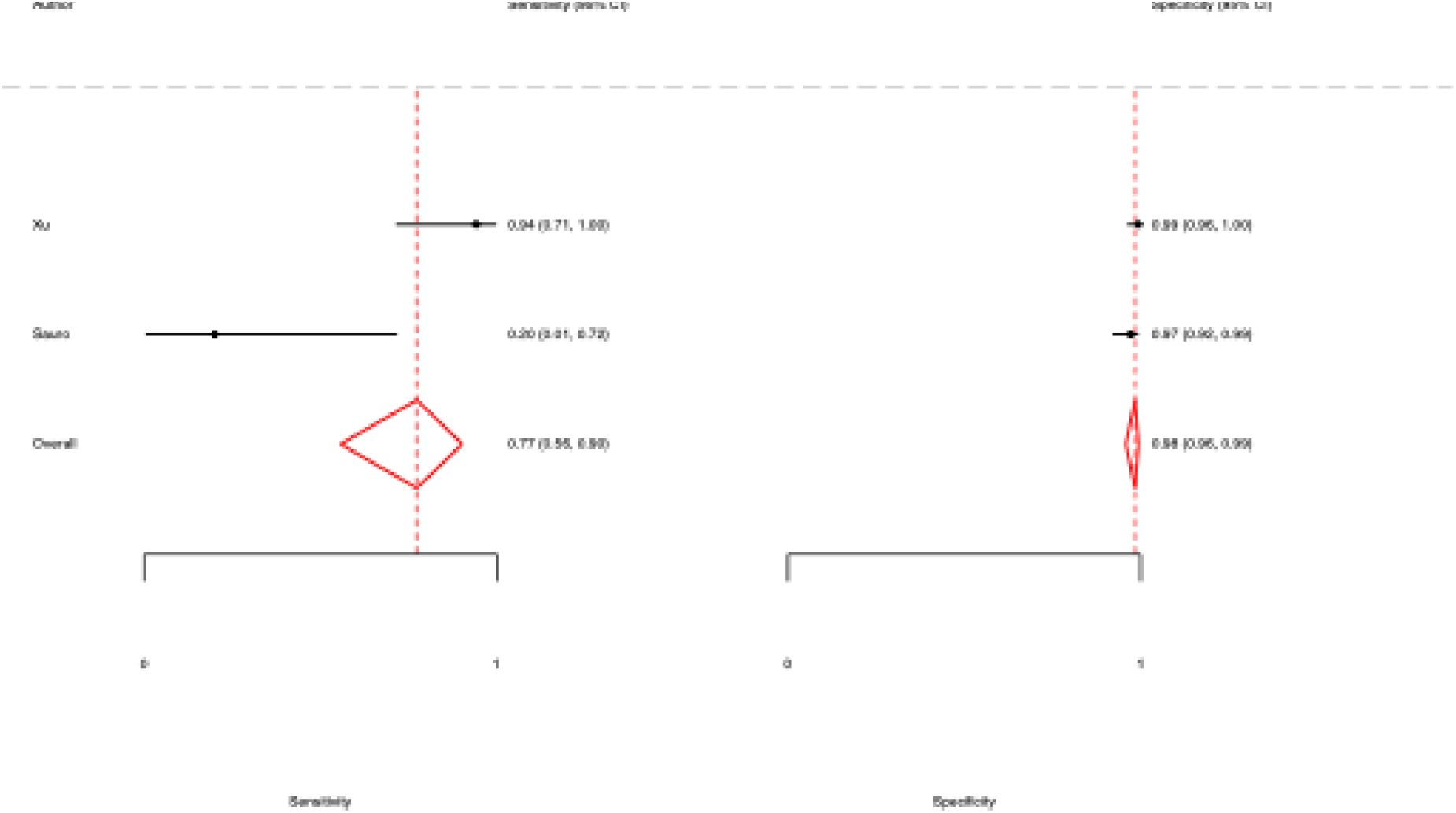
Forest plot of meta-analysis of measures of diagnostic accuracy for reoperations. *Notes:* Only Xu and Sauro are included in meta-analysis as Pache did not provide enough data in their manuscript and supplemental files to reconstruct the necessary two-by-two tables. The metadta command in Stata was used rather than metandi as metandi requires at least four studies. Fixed-effects model used as n < 3. Forest plots in Figures 2 and 3 only generated for reoperations and complications as these variables were common between Xu and Sauro’s validation studies.

## DISCUSSION

The present study found moderate agreement between chart review and the EIAS database, which suggested more accurate and complete data from trained, experienced nurses. There were variations in the accuracy and missingness by site and reviewer, but when compared to the accuracy across other studies (centres) there were no significant differences.

Large administrative and clinical databases such as EIAS are becoming routinely used for epidemiologic studies and also studies aimed at informing clinical decisions (developing and validating clinical decision support tools) and prognosticating outcomes (predicting who will develop complications or those at risk of mortality) in surgery. Leaders in surgical research have advocated for improved rigour in the use of these datasets (e.g., National Surgical Quality Improvement Program or NSQIP, and Veterans Affairs Surgical Quality Improvement Program).(9, 10) There have been calls for a multipronged approach to improve the quality of studies that leverage such databases; ensuring the right database is used for the right research question and that the data within that database is valid and complete.(10) Aligned with this call, the present study was designed to examine the validity of the EIAS database. The consequence of leveraging these databases without considering and reporting the quality of the data and the rigour of the methods used is dire. For example, questionable and contradictory findings, even from the same data, can lead to poor quality of care and outcomes for patients if clinical care is based on the findings.(11) Given the growth of the EIAS and its widespread adoption, it is critically important to validate the data, not just locally but across centres.(3) Our findings, especially when taken into context of other studies,(4, 5) suggest that EIAS is an accurate and complete database. The findings of this study and the others included in our meta-analysis provide support for the use of EIAS to fill the important role of an audit and feedback system for ERAS guidelines, and to conduct studies exploring outcomes related to ERAS surgeries.

This study identified variation in accuracy and completeness by site and by reviewer, raising questions about consistency in data collection and abstraction. EIAS has standardized definitions for each data element and training for data abstractors. In Alberta, nurses with experience and expertise in surgical care and ERAS guidelines abstracted the data using these criteria, as outlined for EIAS. Indeed, we found that the trained nurses did a superior job than our reviewers at abstracting accurate data. To this end, two of our reviewers who were trained physicians had more accurate and complete data abstraction than the non-clinical reviewers. Taken together, this suggests that training and experience with surgical care is critical to high-quality data. Recently there have been developments in the use of natural language processing to abstract data from electronic medical records for ERAS auditing systems. This work is exciting and has the potential to increase the sustainability of ERAS audit and feedback systems by minimizing the resources required to support audit and feedback. However, it is important that these approaches are validated before widespread use.

### Limitations

There are two primary limitations with this study. First, due to constraints surrounding data availability, it was not possible to ascertain all patients who were eligible for ERAS-guided surgeries in Alberta during the study period and therefore it was not possible to determine the coverage of the EIAS database in Alberta. Secondly, an extensive, systematic review was not conducted to identify all possible validation studies for EIAS. However, through the extensive expertise with ERAS evidence and a network of ERAS researchers we are confident that the two studies included in the meta-analysis represent the existing validation studies.

## Data Availability

Deidentified data can be made available upon reasonable request to the corresponding author.

