## Supplemental Files for "Validation of the Enhanced Recovery After Surgery (ERAS) database in Alberta, Canada"

**Supplemental File 1.** Data Abstraction Form

Patient ID: __________

Reviewer: ____________

Date data abstracted: MM / DD / YYYY

**Hospital admission information** ----------------------------------------------------------------------

Hospital admission date: MM / DD / YYYY

Hospital discharge date: MM / DD / YYYY

Type of surgery: _____________

Date of index surgery: MM / DD / YYYY

**Length of stay** ---------------------------------------------------------------------------------------------

Length of stay (days):

Was length of stay missing from EIAS? ☐Yes ☐No

Did length of stay differ in EIAS? ☐Yes ☐No
If yes, describe: ___________________________________________________________

**Complications** ---------------------------------------------------------------------------------------------

Did patient experience a complication related to their surgery? ☐Yes ☐No
If yes, describe nature of complication:
_____________________________________________________________________________
_____________________________________________________________________________
_____________________________________________________________________________

If yes, please categorize complication:
☐Cardiac
☐Respiratory
☐Renal
☐Neurological

☐Gastrointestinal
☐Bleeding
☐DVT/PE
☐Surgical site infection
☐Drug-related
☐Anesthesia-related

If yes, please determine severity (check one):

| | Grade | Definition |
|---|---|---|
| | Grade I | Any deviation from the normal postoperative course without the need for pharmacological treatment or surgical, endoscopic, and radiological interventions. Allowed therapeutic regimens are: drugs as antiemetics, antipyretics, analgetic, diuretics and electrolytes and physiotherapy. This grade also includes wound infections opened at the bedside. |
| | Grade II | Requiring pharmacological treatment with drugs other than such allowed for grade I complications. Blood transfusions and total parenteral nutrition are also included. |
| | Grade IIIa | Requiring surgical, endoscopic, or radiological intervention. Intervention not under general anesthesia. |
| | Grade IIIb | Requiring surgical, endoscopic, or radiological intervention. Intervention under general anesthesia |
| | Grade IVa | Life-threatening complication (including CNS complications) requiring IC/ICU-management. Single organ dysfunction (including dialysis). |
| | Grade IVb | Life-threatening complication (including CNS complications) requiring IC/ICU-management. Multiorgan dysfunction. |
| | Grade V | Death of a patient |

Was complication missing from EIAS? ☐Yes ☐No

Did complication data differ in EIAS? ☐Yes ☐No
If yes, describe: ____________________________________________________________

**Readmission** ------------------------------------------------------------------------------------------------

Hospital readmission: ☐Yes ☐No
If yes, date of subsequent admission: MM / DD / YYYY

Was readmission missing from EIAS? ☐Yes ☐No

Did readmission differ in EIAS? ☐Yes ☐No

If yes, describe: ____________________________________________________________

**Reoperation** ---------------------------------------------------------------------------------------------

Reoperation: ☐Yes ☐No
If yes, date of subsequent surgery: MM / DD / YYYY

Was reoperation missing from EIAS? ☐Yes ☐No

Did reoperation differ in EIAS? ☐Yes ☐No
If yes, describe: ____________________________________________________________

**Hospital mortality** ---------------------------------------------------------------------------------------

Did hospital admission result in death? ☐Yes ☐No

Was hospital mortality missing from EIAS? ☐Yes ☐No

Did hospital mortalityy differ in EIAS? ☐Yes ☐No
If yes, describe: ____________________________________________________________

**Compliance** ----------------------------------------------------------------------------------------------

Was care provided compliant with ERAS?

| Yes | Component | Define ERAS recommended care* |
|---|---|---|
| | Preoperative counselling | |
| | Smoking & alcohol cessation counselling | |
| | Preoperative anemia | |
| | Preoperative nutrition (inc. carbohydrate loading) | |
| | Preoperative bowl prep avoidance | |
| | Interoperative antibiotics | |
| | Interoperative normothermia | |
| | Intraoperative euvolemia | |
| | Postoperative VTE prophylaxis | |
| | Multimodal analgesia | |
| | Nausea control | |
| | Avoid nasogastric suction and intraabdominal drains | |

| | |
|---|---|
| | Postoperative euvolemia |
| | Active mobilization |
| | Removal of urinary catheter |
| | Early feeding with high protein diet |

*Will differ for each type of surgery

Was compliance data missing from EIAS? ☐Yes ☐No

Did compliance data differ in EIAS? ☐Yes ☐No

If yes, describe in detail:

______________________________________________________________________________

______________________________________________________________________________

______________________________________________________________________________

______________________________________________________________________________

**Supplemental File 2:** Agreement by surgery type.

| | Colorectal | Breast | Head & Neck | Pancreas | Urology |
|---|---|---|---|---|---|
| **Patient Outcomes** | | | | | |
| Re-operations | 25.00 | | | | |
| Re-admissions | 84.48 | 81.82 | 75.00 | 100.00 | 100.00 |
| Complications | 70.00 | 70.00 | 62.50 | 44.44 | 42.86 |
| **ERAS Elements** | | | | | |
| Preoperative Anemia | 90.48 | 100.00 | 100.00 | 100.00 | 87.50 |
| Preoperative Nutrition | 57.14 | 9.09 | 11.11 | 44.44 | 50.00 |
| Oral Bowel Preparation | 61.90 | 90.91 | 100.00 | 0.00 | 0.00 |
| Intraoperative Antibiotics | 79.37 | 81.82 | 33.33 | 77.78 | 87.50 |
| Intraoperative Normothermia | 76.19 | 100.00 | 100.00 | 100.00 | 50.00 |
| PONV Prophylaxis | 87.30 | 27.27 | 77.78 | 44.44 | 12.50 |
| Nasogastric Tube | 68.25 | 54.55 | 77.78 | 33.33 | 75.00 |
| VTE Prophylaxis | 91.80 | 9.09 | 44.44 | 88.89 | 87.50 |
| Termination of Urinary Drainage | 79.69 | 81.82 | 77.78 | 100.00 | 100.00 |
| Postoperative Euvolemia | 67.74 | 81.82 | 33.33 | 77.78 | 75.00 |
| Early Mobilization | 95.31 | 100.00 | 100.00 | 100.00 | 100.00 |
| Early Feeding | 66.13 | 90.91 | 100.00 | 100.00 | 100.00 |
| **Overall (Mean)** | 73.19 | 69.94 | 70.93 | 72.22 | 69.13 |

*Abbreviations:* TP = true positive; FN = false negative; FP = false positive; TN=true negative; PONV = postoperative nausea and vomiting; VTE = venous thromboembolism.
